# Neurodevelopmental effects of genetic frontotemporal dementia mutations revealed by total intracranial volume differences

**DOI:** 10.1101/2025.03.28.25324785

**Authors:** Isis So, Arabella Bouzigues, Lucy L. Russell, Phoebe H. Foster, Eve Ferry-Bolder, John C. van Swieten, Lize C. Jiskoot, Harro Seelaar, Raquel Sanchez-Valle, Robert Laforce, Caroline Graff, Daniela Galimberti, Rik Vandenberghe, Alexandre de Mendonça, Pietro Tiraboschi, Isabel Santana, Alexander Gerhard, Johannes Levin, Sandro Sorbi, Markus Otto, Florence Pasquier, Simon Ducharme, Chris R. Butler, Isabelle Le Ber, Maria Carmela Tartaglia, Mario Masellis, James B. Rowe, Matthis Synofzik, Fermin Moreno, Barbara Borroni, Tyler Kolander, Carly Mester, Danielle Brushaber, Kejal Kantarci, Hilary W. Heuer, Leah K. Forsberg, Jonathan D. Rohrer, Bradley F. Boeve, Adam L. Boxer, Howard J. Rosen, Elizabeth C. Finger, Frontotemporal Dementia Prevention Initiative (FPI) Investigators

**Affiliations:** Neuroscience, Schulich School of Medicine & Dentistry, University of Western Ontario, London, ON, Canada; Lawson Health Research Institute, Parkwood Institute, London, ON, Canada; Dementia Research Centre, Department of Neurodegenerative Disease, UCL Queen Square Institute of Neurology, University College London, London, United Kingdom; Department of Neurology and Alzheimer Centre, Erasmus MC University Medical Centre, Rotterdam, the Netherlands; Alzheimer’s disease and Other Cognitive Disorders Unit, Neurology Service, Hospital Clínic, Institut d’Investigacións Biomèdiques August Pi I Sunyer, University of Barcelona, Barcelona, Spain; Clinique Interdisciplinaire de Mémoire, Département des Sciences Neurologiques, CHU de Québec, and Faculté de Médecine, Université Laval, Québec City, QC, Canada; Department of Neurobiology, Care Sciences and Society, Center for Alzheimer Research, Division of Neurogeriatrics, Bioclinicum, Karolinska Institutet, Solna, Sweden; Fondazione Ca’ Granda, IRCCS Ospedale Policlinico, Milan, Italy; University of Milan, Milan, Italy; Laboratory for Cognitive Neurology, Department of Neurosciences, KU Leuven, Leuven, Belgium; Faculty of Medicine, University of Lisbon, Lisbon, Portugal; Fondazione IRCCS Istituto Neurologico Carlo Besta, Milan, Italy; University Hospital of Coimbra (HUC), Neurology Service, Faculty of Medicine, University of Coimbra; Division of Psychology Communication and Human Neuroscience, Wolfson Molecular Imaging Centre, University of Manchester; Department of Neurology, Ludwig-Maximilians Universität München, Munich, Germany; Department of Neuropharmacology, University of Florence, Florence, Italy; Department of Neurology, University of Ulm, Ulm, Germany; University of Lille, Lille, France; Douglas Mental Health University Health Centre, Department of Psychiatry, McGill University; Global Brain Health, Department of Brain Sciences, Imperial College London, UK; Sorbonne Université, Paris Brain Institute – Institut du Cerveau – ICM, Inserm U1127, CNRS UMR 7225, AP-HP - Hôpital Pitié-Salpêtrière, Paris, France; Tanz Centre for Research in Neurodegenerative Diseases, Division of Neurology, University of Toronto, Toronto, ON, Canada; Division of Neurology, Department of Medicine, Sunnybrook Health Sciences Centre; Hurvitz Brain Sciences Program, Sunnybrook Research Institute, University of Toronto, Toronto, ON, Canada; Department of Clinical Neurosciences and Cambridge University Hospitals NHS Trust and Medical Research Council Cognition and Brain Sciences Unit, University of Cambridge, Cambridge, UK; Department of Neurodegenerative Diseases, Hertie-Institute for Clinical Brain Research and Center of Neurology, University of Tübingen; Cognitive Disorders Unit, Department of Neurology, Hospital Universitario Donostia; Neurology Unit, Department of Clinical and Experimental Sciences, University of Brescia; Department of Neurology, Mayo Clinic, Rochester, MN, USA; Department of Quantitative Health Sciences, Mayo Clinic, Rochester, MN, USA; Department of Radiology, Division of Neuroradiology, Mayo Clinic, Rochester, MN, USA; Department of Neurology, Memory and Aging Center, Weill Institute for Neurosciences, University of California, San Francisco, San Francisco, CA, USA; Department of Clinical Neurological Sciences, Schulich School of Medicine & Dentistry, University of Western Ontario, London, ON, Canada

**Keywords:** Frontotemporal dementia, neurodevelopment, hereditary dementia, GRN, MAPT, C9orf72, MRI, total intracranial volume

## Abstract

**Background and Objectives:** Converging evidence hints at neurodevelopmental effects in people at risk of genetic frontotemporal dementia (FTD), including associations between FTD-causing mutations and neurodevelopmental disorders, and differences in young adult mutation carriers compared to familial non-mutation carriers in total intracranial volume (TIV) and cognition. We aimed to investigate TIV and educational attainment differences between adult mutation carriers and familial non-mutation carriers, as measures of the structural and functional neurodevelopmental effects of the FTD-causing genetic mutations.

**Methods:** This cross-sectional cohort study was facilitated through the FTD Prevention Initiative (FPI). Participants, aged 18 to 86 years, were pathogenic mutation carriers of *GRN*, *MAPT*, or *C9orf72*, or familial non-carriers. ANCOVAs were computed per gene to compare outcome means for the main effect of group by carrier status, while controlling for birth decade, sex, and visit site (to account for unique scanners and educational systems). Pearson’s correlations were used to examine associations between TIV and education.

**Results:** Nine-hundred two mutation carriers (mean±SD; age=50.0±13.2 years, sex=55% female, *n*(*GRN*)=298, n(*MAPT*)=187, *n*(*C9orf72*)=417) were compared to 532 familial non-carriers (age=48.0±12.9 years, sex=58% female, *n*(*GRN*)=201, n(*MAPT*)=114), *n*(*C9orf72*)=217). Consistent with prior findings in young adults, *GRN* carriers showed larger TIV compared to familial non-carriers (95% confidence interval [CI]=1431994-1457123, *p*=0.049, *η*^2^*p*=0.008). Larger TIV correlated with higher years of education in *GRN* carriers (95% CI=0.01-0.24, *r*(295)=0.12, *p*=0.03) and *GRN* non-carriers (95% CI=0.08-0.34, r(198)=0.21, *p*=0.002). *MAPT* carriers demonstrated smaller TIV than non-carriers (95% CI=1417819-1450628, *p*=0.039, *η*^2^*p*=0.02). Models with *C9orf72* and education as outcome variables did not reveal significant differences.

**Discussion:** In support of the neurodevelopmental hypothesis of FTD, *GRN* and *MAPT* mutations are linked to likely structural neurodevelopmental changes in TIV, some of which correlate to years of education. These findings motivate further research to identify mechanisms by which FTD mutations influence neurodevelopment and ascertain their suitability as targets for interventions.

## INTRODUCTION

Frontotemporal dementia (FTD) is a common cause of early-onset dementia, with most individuals being clinically diagnosed before 75 years of age.^1^ Its heterogenous and progressive symptomology, affecting behaviour, language, and motor function, carries a devastating burden for patients and caregivers alike.^1,2^ Genetic FTD makes up approximately 20-30% of all FTD cases, with the most common autosomal dominant mutations in one of three genes: chromosome 9 open reading frame 72 (*C9orf72*), microtubule-associated protein tau (*MAPT*), and progranulin (*GRN*).^2–5^

FTD is widely recognized as a neurodegenerative disease, but various studies provide reason to suggest that there are neurodevelopmental effects of genetic FTD. For instance, the most commonly affected genes in FTD are highly penetrant and play crucial roles during neurodevelopment. *C9orf72* hexanucleotide repeat expansions disrupt neuronal motility and cause neuronal apoptosis in zebrafish embryos,^6^ and reduce neural stem cell proliferation as well as decrease thalamic and cortical size during mouse neurodevelopment.^7^ *GRN* acts as a neurotropic factor for neurite outgrowth and neuronal differentiation and survival.^8,9^ *MAPT* mutations disrupt microtubule stabilization and axonal transport.^10^ Furthermore, a recent neuroimaging study of young adults from the Genetic FTD Initiative (GENFI) found that compared to their respective young adult non-mutation carriers, on average, *C9orf72* mutation carriers had smaller total brain and thalamic volumes; *MAPT* mutation carriers had larger total intracranial volumes (TIV), higher levels of education, and better performance on tasks of verbal fluency and attention per the digit span forward; and *GRN* carriers had larger TIV and better performance on the digit symbol task than non-carriers.^11^

Neurodevelopmental effects have been reported in related neurodegenerative diseases that have midlife onset, such as genetic Alzheimer’s disease^12,13^ and Huntington’s disease.^14–17^ For example, in Huntington’s disease, mutation carriers of pathogenic-length CAG expansions show abnormal structural, functional, and cognitive brain development compared to non-mutation.^14,15,17^ This includes structural findings of smaller TIV^14^; initial hypertrophy followed by declines in striatal volume in youth aged 6 to 18 years^15^; and hyperconnectivity followed by declines in the striatal-cerebellar circuitry of youth aged 6 to 12 years, suggestive of compensatory mechanisms.^16^ Functionally, improved cognitive performance is observed in young adult mutation carriers, suggestive of neurodevelopmental benefits of the mutant huntingtin protein prior to neurodegeneration in adult years.^17^

Different approaches can be used to identify neurodevelopmental biomarkers of FTD, including neuroimaging. TIV, which includes the volume of all cranial tissues and the surrounding cerebrospinal fluid, can serve as a proxy measure of premorbid brain growth, and can thus be a plausible correlate of neurodevelopment. As the skull stops growing, TIV stabilizes at around late adolescence.^18^ In normative populations, males have larger TIV than females,^19^ and differences in historical year of birth, as captured by generations and likely reflecting different secular growth rates, are recognized to affect TIV measurement.^20^ TIV has been employed in schizophrenia,^21^ autism spectrum disorder,^22^ and Huntington’s^23^ to examine the neurodevelopmental effects of disease.

Collectively, there are growing hints of neurodevelopmental effects in genetic FTD, though the hypothesis is not yet proven.^11,24–28^ To date, only a few studies on neurodevelopment in FTD have been conducted, typically with small sample sizes, and the field lacks data on youth mutation carriers and non-carriers. TIV differences were found between mutation carriers and non-carriers in young adults between the ages of 18 and 29 from the GENFI cohort, with some beneficial cognitive associations.^11^ The present study extends the examination of previous findings, with an overarching aim of identifying whether markers of structural and functional neurodevelopmental consequences of the *C9orf72*, *MAPT*, and *GRN* genes can be detected in adult mutation carriers. The objectives were to examine whether and how TIV and years of education differ between gene mutation carriers and familial non-carriers, as measures of the potential structural and functional neurodevelopmental outcomes of genetic FTD.

## METHODS

### Participants

This study was facilitated through the FTD Prevention Initiative (FPI). It comprised 902 adult carriers of known pathogenic mutations in *C9orf72* (greater than 30 repeats), *GRN*, or *MAPT*, and 532 familial non-mutation carriers (Table 1 and eTable 1). All participants were enrolled in one of two consortia: the Genetic Frontotemporal Dementia Initiative (GENFI; see eReferences e1), or the ALLFTD consortium (e2), which recently combined the Advancing Research and Treatment for Frontotemporal Lobar Degeneration study (ARTFL; e3) and the Longitudinal Frontotemporal Lobar Degeneration study (LEFFTDS; e4). GENFI phase 1 and 2 included 26 clinical research centers across Europe and Canada, while ALLFTD included 18 clinical research centers across the United States and Canada.

**Table 1.** Demographic characteristics of all participants.

| Characteristic | All |  |  | GRN |  |  | MAPT |  |  | C9orf72 |  |  |
| --- | --- | --- | --- | --- | --- | --- | --- | --- | --- | --- | --- | --- |
|  | Total | Carriers | Non-carriers | Carriers | Non-carriers | Between group comparison | Carriers | Non-carriers | Between group comparison | Carriers | Non-carriers | Between group comparison |
| Sample size (*n*) | 1434 | 902 | 532 | 298 | 201 | NA | 187 | 114 | NA | 417 | 217 | NA |
| Age, years (mean ± SD) | 50.0 ± 13.2 | 51.2 ± 13.2 | 48.0 ± 12.9 | 53.8 ± 12.4 | 50.9 ± 13.1 | $t=2.46$, $p=0.01^*$, $d=0.22$ | 46.0 ± 12.6 | 43.9 ± 12.7 | $t=1.34$, $p=0.18$, $d=0.16$ | 51.8 ± 13.4 | 47.4 ± 12.1 | $t=4.01$, $p=6.79e-5^{***}$, $d=0.34$ |
| Birth Decade (*n*) |  |  |  |  |  |  |  |  |  |  |  |  |
| 1920-1929 | 2 | 0 | 2 | 0 | 2 |  | 0 | 0 |  | 0 | 0 |  |
| 1930-1939 | 16 | 10 | 6 | 4 | 4 |  | 0 | 0 |  | 6 | 2 |  |
| 1940-1949 | 147 | 105 | 42 | 43 | 17 |  | 10 | 7 |  | 52 | 18 |  |
| 1950-1959 | 305 | 208 | 97 | 82 | 50 | $\chi^2=11.7$, $p=0.11$, $d=-0.17$ | 31 | 12 | $\chi^2=4.44$, $p=0.62$, $d=-0.14$ | 95 | 35 | $\chi^2=23.3$, $p=7.00e-4^{***}$, $d=-0.27$ |
| 1960-1969 | 321 | 218 | 103 | 76 | 45 |  | 44 | 21 |  | 98 | 37 |  |
| 1970-1979 | 366 | 193 | 173 | 59 | 55 |  | 51 | 40 |  | 83 | 78 |  |
| 1980-1989 | 226 | 139 | 87 | 29 | 22 |  | 39 | 26 |  | 71 | 39 |  |
| 1990-1999 | 49 | 28 | 21 | 5 | 6 |  | 11 | 7 |  | 12 | 8 |  |
| 2000-2009 | 2 | 1 | 1 | 0 | 0 |  | 1 | 1 |  | 0 | 0 |  |
| Sex (*n*) female: male | 803:631 | 495:407 | 308:224 | 172:126 | 113:88 | $\chi^2=0.06$, $p=0.81$, $d=-0.03$ | 105:82 | 61:53 | $\chi^2=0.11$, $p=0.74$, $d=-0.05$ | 218:199 | 134:83 | $\chi^2=4.81$, $p=0.03^*$, $d=0.19$ |
| Education, years (mean ± SD) | 14.6 ± 4.0 | 14.5 ± 4.4 | 14.8 ± 3.2 | 14.3 ± 3.8 | 14.7 ± 3.7 | NA | 15.0 ± 6.9 | 14.9 ± 2.8 | NA | 14.4 ± 3.2 | 14.9 ± 3.0 | NA |
| Handedness (*n*) |  |  |  |  |  |  |  |  |  |  |  |  |
| Right | 1300 | 811 | 489 | 269 | 193 | $\chi^2=6.48$, $p=0.09$, $d=-0.05$ | 173 | 100 | $\chi^2=1.96$, $p=0.38$, $d=0.02$ | 369 | 196 | $\chi^2=7.52$, $p=0.11$, $d=-0.16$ |
| Left | 110 | 76 | 34 | 24 | 6 |  | 14 | 14 |  | 38 | 14 |  |
| Ambidextrous | 22 | 13 | 9 | 5 | 2 |  | 0 | 0 |  | 8 | 7 |  |
| Unknown | 1 | 1 | 0 | 0 | 0 |  | 0 | 0 |  | 1 | 0 |  |
| Blinded Sites (*n*) | 45 | 43 | 36 | 41 | 33 | NA | 27 | 18 | NA | 41 | 33 | NA |

\* $p<0.05$ , \*\*\* $p<0.001$ between carriers and non-carriers. Blinded sites (*n*) refer to the number of sites which saw the specified group of participants. An independent t-test was not performed between groups for variable of education, since this was a planned secondary outcome that would be analyzed in greater detail. Race was reported for the GENFI study but not for the ALLFTD study, hence there is no aggregated information to report for the primary analysis. Abbreviations: SD, standard deviation; NA, not applicable.

Ninety-three young adult GENFI participants who were included in the previously published TIV analysis on GENFI young adults aged 18-29 years^11^ were excluded. The data analyzed in this study includes participants from the following: GENFI Data Freeze 6 from Phase 1 (GENFI1, 2012-15) and Phase 2 (GENFI2, 2015-19), and ALLFTD Data Freeze 13, ARTFL/LEFFTDS from 2015-2020, and ALLFTD from 2020-2023.

### Standard Protocol Approvals, Registrations, and Patient Consents

Local ethics committees at each of the sites approved the study, and all participants or their proxy decision maker provided written informed consent, in accordance with the Declaration of Helsinki.

### Study Design and Procedures

The GENFI and ALLFTD investigations are prospective and longitudinal in design, and collect demographic, neuroimaging, neuropsychological, behavioural, and clinical outcomes. Given the general stability of TIV and education level past young adulthood, baseline demographic and neuroimaging data were analyzed in this cross-sectional cohort study. Both the GENFI and ALLFTD cohorts contain symptomatic *C9orf72*, *MAPT*, or *GRN* mutation carriers, pre-symptomatic mutation carriers, and familial non-mutation carriers. Since the genes of interest in this study are highly penetrant, symptomatic and pre-symptomatic mutation carriers were considered as one group (carriers), and familial non-mutation carriers were considered the comparator (non-carriers).

### Neuroimaging

GENFI and ALLFTD image acquisition and preprocessing protocols have been delineated elsewhere.^11,29–31^ Pertinent to this study, GENFI participants underwent T1-weighted MRI based on the GENFI protocol using a 1.5 T (Siemens, GE) or 3T scanner (Siemens Trio, Siemens Skyra, Siemens Prisma, Philips Achieva, GE Discovery MR750). The sequence parameters were: 256 × 256 × 208 matrix; 208 slices; 1.1 mm isotropic voxel size; flip angle of 8°; and echo time and repetition time varied by vendor. ALLFTD participants underwent T1-weighted MRI using 3T scanners (model information reported elsewhere)^30^. Magnetization Prepared Rapid Gradient Echo images were obtained using these parameters: 240 × 256 × 256 matrix; 170 slices; voxel size = 1.05 × 1.05 × 1.25 mm3; flip angle, echo time and repetition time varied by vendor.

A standard imaging protocol was used across all centers, managed, reviewed for quality, and preprocessed by the respective core imaging groups per consortia. Prior to preprocessing, all images were visually inspected for quality control, and those with excessive movements or image artifacts were removed. The preprocessing steps have been previously reported,^30,31^ and include bias field correction with the N3 algorithm, segmentation using SPM12 v6470 (Statistical Parametric Mapping, Wellcome Trust Centre for Neuroimaging, London, UK), normalization to a customized group template (generated using the Large Deformation Diffeomorphic Metric Mapping framework), and spatial smoothing with an 8 mm full width half maximum Gaussian kernel. TIV, which includes all gray matter, white matter, and CSF, was computed with SPM12 v6470 running under MATLAB R2014b AQ33 (MathWorks, Natick, MA, USA).

### Statistical Analysis

All analyses were computed in R v3.6.3. Demographic comparisons between carriers and non-carriers were conducted using t-tests (age at visit) or chi-square tests (sex, birth decade, race, handedness). Statistical assumptions of data normality were determined using histograms, Q-Q plots, and the Shapiro-Wilk test (p>0.05). Analysis of covariance (ANCOVA) assumptions of homogeneity of regression slopes, and independence of the covariates and independent variable, were also examined and passed before model computation. ANCOVAs were used to compare outcome means for the main effect of group (carriers vs. non-carriers), with separate models for the three gene groups (*C9orf72*, *MAPT*, *GRN*), while controlling for three covariates of non-interest: birth decade, sex, and visit site. Visit site was used as it accounted for unique scanners per site, as well partial accounting of variance in educational systems related to cultural differences per country and region. GENFI and ALLFTD participants were analyzed together in primary analyses (see eAppendix for findings of separate cohort analysis). Outliers for both TIV and education were determined with all participant data in aggregate per gene; a cut-off of greater than 3 SD was used. Any data point classified as an outlier was removed prior to analysis.

Birth decade was included as a covariate rather than age at time of visit, as skull changes and TIV are understood to be stable from late adolescence,^18^ and changes in secular growth rates have been recognized to have a measurable influence on TIV changes over different generations.^20^ This approach of categorizing birth by decade to control for the generational effect on changes in secular growth rate has similarly been used by others.^32^ In our analysis, birth decade was determined by the year in which a participant was born; it ranged from the beginning of a decade to ten years after (e.g., 1930 to 1939, inclusive), and included seven time periods: 1930s, 1940s, 1950s, 1960s, 1970s, 1980s, and 1990s.

While there were no predictions of differential sex effects of the FTD mutations, there are known sex-specific differences in TIV^19^ and education.^33^ Therefore, in the primary analysis, the outcome variables of interest were analyzed with a sex-aggregated approach (males and females combined), with sex as a covariate. Planned follow-up sensitivity analyses used a sex-stratified approach, where males and females were analyzed separately. Sensitivity analysis for site was also conducted, censoring sites that had only one participant (i.e., only one carrier or non-carrier): 4/31 sites for *GRN*, 7/28 sites for *MAPT*, and 9/42 sites for *C9orf72* contained only one participant.

To determine whether brain size, particularly if larger, reflects the usual positive relationship between TIV and education, per gene, Pearson’s correlations were performed to examine potential associations between TIV and education for carriers and non-carriers.

### Exploratory *MAPT* genetic mutation subtype analysis

To assess for potential differential effects of different *MAPT* mutations, a separate model was computed for *MAPT* participants with mutation group as an additional covariate. *MAPT* mutations were categorized into 1 of 5 groups by their underlying pathophysiology and/or functional consequences, as defined previously (eTable 2).^34^

### Data Availability

Anonymized data may be requested from the GENFI and ALLFTD projects (available in eReferences), though certain elements of the data from both consortia may be restricted to protect the confidentiality of participants. Analytic R code can be made available upon request.

## RESULTS

### Participants

The primary analyses of the combined cohort examined 1434 participants, of whom 902 were mutation carriers and 532 were non-carriers (Table 1). Participants were categorized based on genetic mutations, resulting in 298 *GRN* carriers, 201 *GRN* familial non-carriers, 187 *MAPT* carriers, 114 *MAPT* familial non-carriers, 417 carriers of the *C9orf72* repeat expansion, and 217 non-carrier relatives of *C9orf72.* The mean age of all participants at their baseline visit, at which TIV was measured and years of education were assessed, was 50.0 years (SD=13.2, range=18 to 89.7).

*GRN* carriers were slightly older than their familial non-carriers (53.8 vs. 50.9 years, *p*=0.01, *d*=0.22). There were no differences in birth decade, sex, and handedness, between *GRN* carriers and non-carriers (*p*>0.05). No differences were observed in *MAPT* carriers and non-carriers in any of these demographic variables (*p*>0.05). *C9orf72* repeat expansion carriers differed from their familial non-carriers in age (*p*=6.79e-5, *d*=0.34), birth decade (*p*=7.00e-4, *d*=-0.27), and sex (*p*=0.03, *d*=0.19), with carriers being older, born in earlier decades, and having a higher ratio of males; however, these differences were all small in effect size (*d*<0.5). No differences were observed between *C9orf72* carriers and non-carriers in handedness (*p*>0.05).

### GRN

#### TIV

*GRN* carriers exhibited larger TIV than familial non-carriers: *F*(1,495)=3.89, 95% confidence interval (CI)=85.4-40977, *p*=0.049, *η*^2^*p*=0.008 (Figure 1a). As predicted, birth decade, sex, and site were associated with TIV, with later birth decades (*F*(7,495)=2.15, *p*=0.04, *η*^2^*p*=0.03) and males (*F*(1,495)=348, *p*<2.2e-16, *η*^2^*p*=0.43) presenting with greater TIV. There were no differences or interactions with sex in the sex-aggregated sensitivity analysis. Findings from the site sensitivity analysis, where sites containing only one participant was removed, were consistent with that of the primary analysis (eAppendix; see eTable 3 for outcome variable means before and after covariate adjustments).

**Figure 1.**
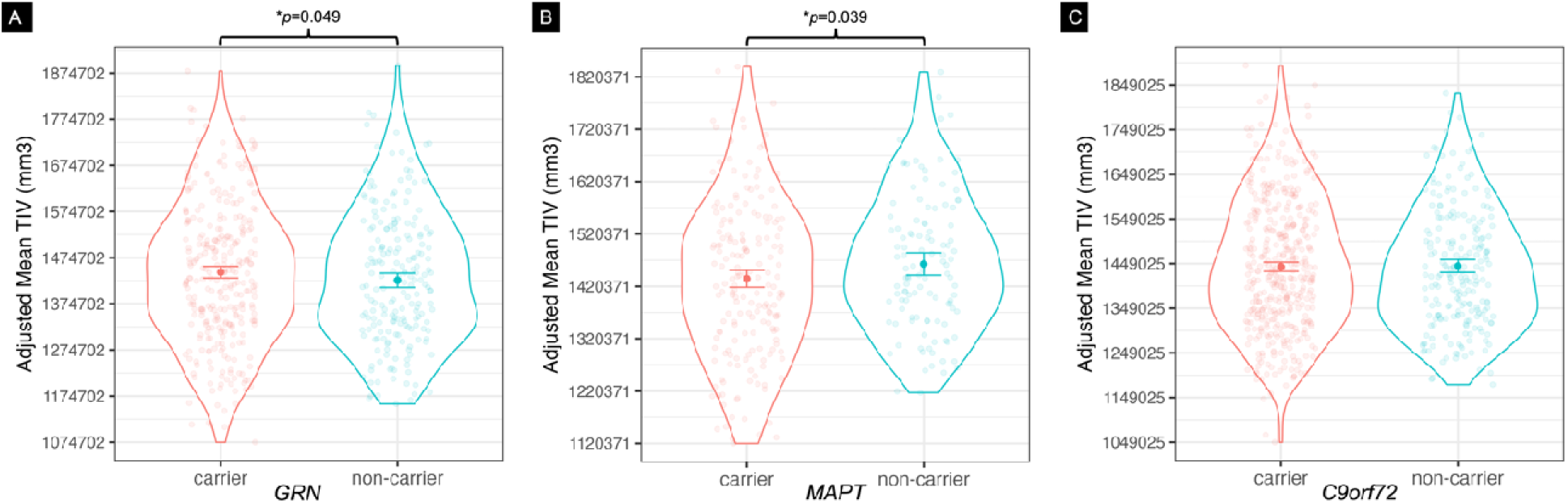
Mean total intracranial volume (TIV) for carriers and non-carrei rs of FTD-causing genetic mutations. TIV measured in mm^3^, adjusted for differences in birth decade, sex, and visit site. TIV differences were observed in **(A)** *GRN* carriers, who had larger TIV, and **(B)** *MAPT* carriers, who had smaller TIV. **(C)** Carriers of the *C9orf72* hexanucleotide repeat expansion showed no differences in TIV relative to non-carriers. \**p*<0.05.

### Education

*GRN* carriers and non-carriers did not differ in years of education (Figure 2a). Main effects were observed for birth decade (*F*(7,497)=6.09, *p*=8.07e-7,*η*^2^*p*=8.50e-2) and site (*F*(30,497)=5.06, *p*=2.89e-15, *η*^2^*p*=0.25). There were no interactions with carrier status (*b*=49.4, SEM=44.2, *t*(497)=1.12, *p*=0.26), and the pattern of results remained the same in the site sensitivity analyses.

**Figure 2.**
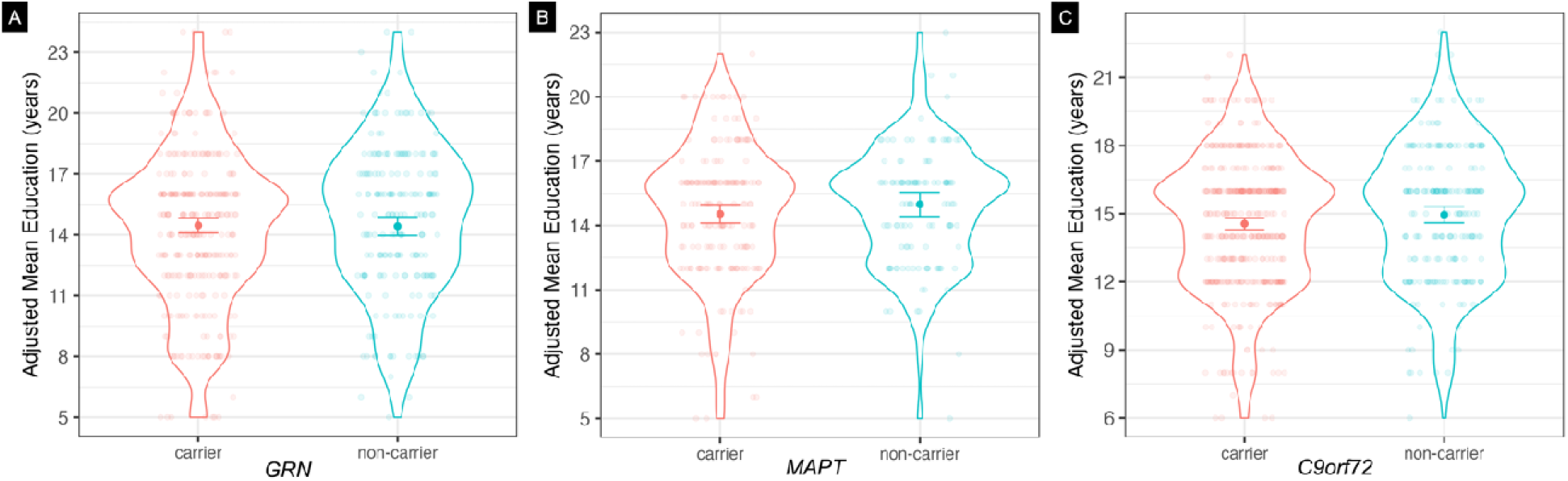
Mean years of education by carriers and non-carriers of FTD-causing genetic mutations. Measured in years, adjusted for differences in birth decade, sex, and visit site. TIV differences were not observed in **(A)** *GRN* carriers, **(B)** *MAPT* carriers, or **(C)** *C9orf72* hexanucleotide repeat expansion carriers compared to respective non-carriers.

### Correlation between TIV and Education

Larger TIV was associated with greater years of education in *GRN* carriers (95% CI=0.01- 0.24, *r*(295)=0.12, *p*=0.03) and non-carriers (95% CI=0.08-0.34, *r*(198)=0.21, *p*=0.002).

### MAPT

#### TIV

*MAPT* carriers had smaller TIV than their familial non-carriers: *F*(1,299)=4.31, 95% CI=- 58248-(-1545), *p*=0.039, *η*^2^*p*=0.02 (Figure 1b). Sex was associated with TIV, with males having larger TIV: *F*(1,299)=192, *p*<2e-16, *η*^2^*p*=0.40. When sex was stratified, female *MAPT* carriers had smaller TIV than non-carriers: *F*(1,163)=4.88, *p*=0.03, *η*^2^*p*=0.04; no significant difference existed between groups in the male *MAPT* model, though the directionality of the means in male *MAPT* carriers and non-carriers matched that of the primary analysis and female only analysis. When *MAPT* mutation group was accounted for as a covariate, there was a trend of a main effect of mutation group: *F*(1,298)=3.63, 95% CI=-58690-(-1943), *p*=0.06, *η*^2^*p*=0.02, but there were no interactions with TIV.

### Education

Participants who carried a *MAPT* mutation did not vary in years of education compared to non-carriers (Figure 2b). The main effect of site was significant in its association with education (*F*(22,298)=1.98, *p*=0.01, *η*^2^*p*=0.18), birth decade approached significance in its association with education (*F*(6,298)=2.06, *p*=0.06, *η*^2^*p*=0.04), and there was a group x birth decade interaction (*b*=2.98, SEM=1.51, *t*(298)=1.98, *p*=0.049); however, upon inspection, there was no consistent pattern of interest.

The removal of sites with only one participant also yielded non-significance, but separate models per GENFI and ALLFTD cohorts were interesting (eTable 4). Analysis of *MAPT* participants in the ALLFTD cohort alone yielded non-significance; however, in *MAPT* participants belonging to the GENFI cohort, the main effect of group approached significance while controlling for birth decade, sex, and site, where *MAPT* carriers tended to have fewer years of education than non-carriers: *F*(1,152)=3.46, *p*=0.07, *η*^2^*p*=0.03. In GENFI participants, birth decade and site also approached significance in its association with years of education: *F*(5,152)=2.13, *p*=0.07, *η*^2^*p*=0.08, and *F*(15,152)=1.56, *p*=0.09, *η*^2^*p*=0.15, respectively. A similar pattern of results was observed when the influence of *MAPT* mutation group was accounted for: *F*(1,152)=3.83, *p*=0.05, *η*^2^*p*=0.03.

### Correlation between TIV and Education

There were no significant correlations between TIV and years of education in *MAPT* carriers or non-carriers. When participants were stratified by sex, the expected trend of larger TIV associated with greater years of education in male *MAPT* carriers was observed: 95% CI=- 0.008-0.41, *r*(78)=0.21, *p*=0.06.

### C9orf72

#### TIV

There was no difference in TIV between repeat expansion carriers and non-carriers (Figure 1c). Main effects of sex and site were present; sex: *F*(1,627)=486.5, *p*<2.2e-16, *η*^2^*p*=0.46, with males having greater TIV than females; and site: *F*(41,627)=2.87, *p*=2.65e-08, *η*^2^*p*=0.17. The sex and site sensitivity analyses showed similar pattern of results (eAppendix).

### Education

No difference was observed in years of education between *C9orf72* repeat expansion carriers and non-carriers (Figure 2c), while main effects of birth decade (*F*(6,620)=2.72, *p*=0.01, *η*^2^*p*=0.03) and site (*F*(41,620)=4.52, *p*<2e-16, *η*^2^*p*=0.25) were observed, with no significant interactions. *C9orf72* participants born in later birth decades were associated with having greater years of education. All sensitivity analyses yielded non-significant findings in similar patterns to the primary analysis.

### Correlation between TIV and Education

Larger TIV was linked to greater years of education in *C9orf72* repeat expansion carriers (95% CI=0.02-0.21, *r*(413)=0.12, *p*=0.02), and approached significance in non-carriers (95% CI=- 0.02-0.25, *r*(213)=0.12, *p*=0.09). When participants were stratified by sex, we observed in male *C9orf72* carriers the expected trend of larger TIV correlated with greater years of education: 95% CI=0.13-0.39, *r*(197)=0.27, *p*=0.0001.

## DISCUSSION

This investigation revealed that a macroscopic neurodevelopment marker, TIV, differed in FTD-associated mutation carriers. Adult carriers of FTD-causing *GRN* mutations have larger TIV than familial non-carriers, while *MAPT* carriers have smaller TIV than their respective non-carriers, both with small effect sizes. Carriers of the *C9orf72* hexanucleotide repeat expansion did not show differences in TIV compared to non-carriers. Our *GRN* and *C9orf72* findings replicated those observed in a smaller young adult sample,^11^ whereas our *MAPT* results were distinct. We did not observe differences in years of education between carriers and non-carriers of any of the genetic mutations of interest. Expected positive correlations between TIV and years of education were present in both *GRN* carriers and non-carriers, and *C9orf72* repeat expansion carriers and non-carriers. The expected influence of sex and birth decade were observed, with males on average having larger TIV and later birth decades associated with larger TIV and greater years of education in all of the gene groups. Collectively, our findings support the hypothesis that some FTD-causing genetic mutations influence structural neurodevelopment.

We identified that adult carriers of *GRN* mutations have greater TIV than familial non-carriers, consistent with prior findings from the GENFI young adult sample.^11^ In FTD, *GRN* mutations are mainly heterozygous loss-of-function mutations that lead to haploinsufficiency in progranulin, a secreted growth factor important for neurodevelopmental processes including neurite outgrowth, and neuronal differentiation and survival.^3,35^ Homozygous *GRN* mutations impair early lysosomal storage function, leading to later neurodegeneration in FTD.^35,36^ Progranulin also promotes synaptic development, as shown in young progranulin knockout mice that had diminished synaptic connectivity, plasticity, and dendritic spine density in the hippocampus,^37^ raising the possibility that loss-of-function *GRN* mutations in FTD could lead to less synaptic pruning and thus potentially greater cortical volumes and resultant larger TIV.

While there were no significant differences in years of education between *GRN* carriers and non-carriers, we observed larger TIV correlated with greater years of education in both *GRN* carriers and non-carriers. Such a correlation was expected in non-carriers as it pertains to the usual pattern observed in healthy adults.^38^ Larger TIV in *GRN* carriers was likewise associated with greater years of education, suggesting that early neurodevelopmental effects related to *GRN* haploinsufficiency are potentially neutral or advantageous with respect to brain function. This theory is supported by findings of enhanced cognitive performance in young adult *GRN* carriers, who performed better in the digit symbol task than non-carriers and performed as well as non-carriers across other cognitive tasks.^11^ Further studies are necessary to determine the driving factors behind the increased TIV in *GRN* mutation carriers.

The current observation of smaller TIV in *MAPT* carriers is in line with literature which supports links between tau and human neurodevelopment.^39–42^ For example, low plasma tau levels have been linked to adolescents with early-onset psychosis, which was correlated with smaller surface area of the orbitofrontal cortex.^39^ Alternative splicing of *MAPT* mRNA transcripts in humans has also been reported to undergo rapid and marked changes between the last trimester of fetal development and the first post-natal months, resulting in differential spatial and temporal expression of tau in the developing brain; these changes coincide with a critical phase of neuronal migration required for establishing mature neuronal connectivity.^40^ In human induced pluripotent stem cell models of FTD, *MAPT* mutations resulted in prolonged maturation of cortical neurons and altered electrophysiological properties, such as depolarized resting membrane potential, increased input resistance, and changes in action potential,^41^ as well as diminished proliferation capacity of neural progenitors and disrupted Wnt/Shh signalling pathways.^42^ Intriguingly, related signalling pathways have been acknowledged for regulating developmental pathways that contribute to TIV: the WNT/ß-catenin pathway is involved in meningeal development, which confers downstream effects on calvarial and brain development^43^; and the Shh-mediated pathways regulate calvarial suture morphogenesis and osteogenesis.^44^ Neurodevelopmental models will be required to determine if *MAPT* mutations causing FTD affect TIV via such direct mesenchymal effects, or indirect effects based on differences in maximum brain volume.

In addition, we explored the effects of *MAPT* mutation type, due to evidence that different *MAPT* mutations confer different functional consequences and underlying pathology, leading to systematic differences in age of FTD onset and eventual death.^34^ Of particular interest, in induced pluripotent stem cells, the *MAPT* IVS10+16 mutation increases 4R tau isoform expression and impairs neuronal progenitor proliferation. Although the effect sizes of the models marginally increased with the addition of *MAPT* mutation group as a covariate, there were no specific mutation type main effects or interactions.

Similar to *GRN*, there were no differences in years of education between *MAPT* carriers and non-carriers in the primary analyses. In a genome-wide association study, *MAPT* was one of the top candidate genes associated with years of education, and gene function analysis revealed *MAPT* mutations during pre-natal stages of development to be associated with impaired dendrite morphogenesis, altered morphology of hippocampal mossy fibers, and atypical axonal guidance,^45^ which are processes involved in long-term potentiation, synaptic plasticity, memory formation, and learning.

The findings of smaller TIV and no difference in years of education in *MAPT* carriers contrast prior findings in GENFI young adults.^11^ Potential reasons for the discrepant findings include a smaller sample size in the young adult study, birth decade or cohort effects. To further explore these possibilities, we replicated those methods, assessing only young adults aged 18 to 29, with age as a covariate instead of birth decade.^11^ In this sub-cohort of 28 young adults, where 27 were from ALLFTD and there was 1 new GENFI participant, TIV was smaller for *MAPT* mutation carriers. Although the distribution of *MAPT* mutation groups was similar between the sub-cohort and the larger cohort, we consider the present findings most robust given the larger sample size, and attribute the differences as likely due to cohort effects of the smaller sample,^11^ including unmeasured differences in socioeconomic status and education systems.

Carriers of the *C9orf72* hexanucleotide repeat expansion did not differ in TIV from non-carriers. This replicates the findings observed in GENFI young adult *C9orf72* carriers (mean age 26 years), which also revealed smaller total brain and thalamic volumes.^11^ However, this does not indicate an absence of neurodevelopmental effects. Less gyrification has been observed in asymptomatic *C9orf72* repeat expansion carriers in parietal, occipital, and temporal regions^28^; since gyrification is a process that begins during third trimester of pregnancy and peaks during childhood^46^, these findings are supportive of neurodevelopmental consequences of *C9orf72* on brain structure, even if TIV is unaffected. Others have highlighted associations between neurodevelopmental disorders and *C9orf72* repeat expansion, such as a higher probability of schizophrenia and autism spectrum disorder in young relatives of *C9orf72* carriers compared with non-carriers.^26^

Various studies support that *C9orf72* affects cellular and molecular processes involved in early stages of neurodevelopment. For instance, *C9orf72* hexanucleotide repeat expansions can disrupt neuronal motility and cause neuronal apoptosis in zebrafish embryos,^6^ and also reduce neural stem cell proliferation and decrease thalamic and cortical size during mice during neurodevelopment.^7^ Indeed, studies suggest that both loss-of-function^4^ and gain-of-function^7^ effects of *C9orf72* repeat expansions contribute to neurodevelopmental effects, similar to the loss- and gain-of-function mechanisms proposed for *C9orf72* induced ALS/FTD neurodegeneration.^47^ These mechanisms, which engage at different times and in different brain regions in mouse neurodevelopment,^48^ could counteract each other, thereby concealing macrostructural neurodevelopmental effects of *C9orf72* on TIV.

*C9orf72* carriers had similar years of education to non-carriers, and a normal positive correlation between TIV and years of education was observed in carriers and trending towards significance in non-carriers. We acknowledge that education levels are widely influenced by socioeconomic and environmental factors unmeasured in this cohort.^49,50^ Future studies including additional demographic variables known to impact educational attainment, as well as prospective cognitive assessments in youth, are needed to better delineate potential neurodevelopmental effects of *C9orf72* on brain function.

While this cohort was large for a rare genetic disease, and enabled greater statistical power than the initial young adult sample,^11^ we acknowledge several limitations. TIV models for *GRN* and *MAPT* had small effect sizes, as did the correlations between TIV and education in *GRN* carriers and non-carriers. Additionally, several potential confounders were not available, such as participant height, which affects cranium size and thus TIV, as well as nutritional and socioeconomic status, which influence years of education. The comparably smaller sample of *MAPT* participants and unequal distribution of participants per *MAPT* mutation group also limited our ability identify potential differential effects of different *MAPT* mutations.

In summary, this study provides insight and evidence supporting the neurodevelopmental hypothesis of FTD, and has several implications for future examination. *GRN* and *MAPT* mutations are associated with changes in TIV, supporting neurodevelopmental effects in carriers of these FTD mutations. In particular, the now replicated finding that *GRN* mutation carriers have larger TIV than non-carriers, and the preserved association between TIV and years of education indicates this genetic variation influences brain structure, and is potentially associated with advantageous functional effects during early development. Studies evaluating the cellular and molecular mechanisms by which these mutations affect development in non-human animal models highlight potential mechanisms by which these FTD mutations may impact neurodevelopment. Future directions investigating these pathways in human genetic FTD models may identify novel treatment targets for maintaining function in genetic FTD beyond young adulthood.

## Supporting information

eAppendix

## ACKNOWLEDGEMENT

The authors thank all GENFI and ALLFTD participants for their contributions to the study.

## STUDY FUNDING

This study was supported by Canadian Institutes of Health Research (CIHR) grants #470797 and #452843, held by E.F., who also holds funding from CIHR grant #327387. I.S. is supported by a BrainsCAN MD/PhD Award, CIHR Canada Graduate Scholarship-Doctoral #193336, and Parkwood Institute Research Cross-Theme Collaboration Studentship (funded by the St. Joseph’s Health Care Foundation). J.C.V.S., L.C.J. and H.S. are supported by the Dioraphte Foundation grant 09-02-03-00, Association for Frontotemporal Dementias Research Grant 2009, Netherlands Organization for Scientific Research grant HCMI 056-13-018, ZonMw Memorabel (Deltaplan Dementie, project number 733 051 042), ZonMw Onderzoeksprogramma Dementie (YOD-INCLUDED, project number10510032120002), EU Joint Programme-Neurodegenerative Disease Research-GENFI-PROX, Alzheimer Nederland and the Bluefield Project. R.S-V. is supported by Alzheimer’s Research UK Clinical Research Training Fellowship (ARUK-CRF2017B-2) and has received funding from Fundació Marató de TV3, Spain (grant no. 20143810). R.S.-V. was funded at the Hospital Clinic de Barcelona by Instituto de Salud Carlos III, Spain (grant code PI20/00448 to RSV) and Fundació Marató TV3, Spain (grant code 20143810 to RSV). RL is supported by the Canadian Institutes of Health Research and theChaire de Recherche sur les Aphasies Primaires Progressives Fondation Famille Lemaire. C.G. received funding from EU Joint Programme-Neurodegenerative Disease Research-Prefrontals Vetenskapsrådet Dnr 529-2014- 7504, EU Joint Programme-Neurodegenerative Disease Research-GENFI-PROX, Vetenskapsrådet 2019-0224, Vetenskapsrådet 2015-02926, Vetenskapsrådet 2018-02754, the Swedish FTD Inititative-Schörling Foundation, Alzheimer Foundation, Brain Foundation, Dementia Foundation and Region Stockholm ALF-project. C.G. is supported by the Swedish Frontotemporal Dementia Initiative Schörling Foundation; Vetenskapsrådet (Swedish Research Council) JPND Prefrontals, 2015–02926, 2018–02754, and JPND-GENFI-PROX 2019-02248; Swedish Alzheimer Foundation, ALF-project Region Stockholm, Karolinska Institutet Doctoral Funding, KI Strat-Neuro, Swedish Dementia Foundation, and Swedish BrainFoundation. R.V. has received funding from the Mady Browaeys Fund for Research into Frontotemporal Dementia. J.L. received funding for this work by the Deutsche Forschungsgemeinschaft German Research Foundation under Germany’s Excellence Strategy within the framework of the Munich Cluster for Systems Neurology (EXC 2145 SyNergy—ID 390857198). M.O. has received funding from Germany’s Federal Ministry of Education and Research (BMBF). S.D. receives salary funding from the Fonds de Recherche du Québec – Santé, and receives funding from the Canada First Research Excellence Fund, awarded to McGill University for the Healthy Brains, Healthy Lives initiative. M.M. was, in part, funded by the UK Medical Research Council, the Italian Ministry of Health and the Canadian Institutes of Health Research as part of a Centres of Excellence in Neurodegeneration grant, and also Canadian Institutes of Health Research operating grants (Grant #s: MOP- 371851 and PJT- 175242) and funding from the Weston Brain Institute. J.B.R. has received funding from the Welcome Trust (103838; 220258) and is supported by the Cambridge University Centre for Frontotemporal Dementia, the Medical Research Council (MC_UU_00030/14; MR/T033371/1) and the National Institute for Health Research Cambridge Biomedical Research Centre (NIHR203312). F.M. is supported by the Tau Consortium and has received funding from the Carlos III Health Institute (PI19/01637). J.D.R. is supported by the Bluefield Project and the National Institute for Health and Care Research University College London Hospitals Biomedical Research Centre, and has received funding from an MRC Clinician Scientist Fellowship (MR/M008525/1) and a Miriam Marks Brain Research UK Senior Fellowship. Several authors of this publication (J.C.V.S., M.S., R.V., A.d.M., M.O., R.V., J.D.R.) are members of the European Reference Network for Rare Neurological Diseases (ERN-RND) - Project ID No 739510. This work was also supported by the EU Joint Programme—Neurodegenerative Disease Research GENFI-PROX grant [2019-02248; to J.D.R., M.O., B.B., C.G., J.C.V.S. and M.S]. L.K.F., B.F.B., and K.K. each receive research support from the NIH. A.L.B. receives research support from the NIH, the Tau Research Consortium, the Association for Frontotemporal Degeneration, Bluefield Project to Cure Frontotemporal Dementia, Corticobasal Degeneration Solutions, the Alzheimer’s Drug Discovery Foundation and the Alzheimer’s Association. H.J.R. receives research support from the NIH and the state of California.

## DISCLOSURE

J.D. Rohrer has received consulting fees from UCB, AC Immune, Astex Pharmaceuticals, Biogen, Takeda, and Eisai and is part of the Medical Advisory Board for Alector, Arkuda Therapeutics, Wave Life Sciences, and Prevail Therapeutics. R. Sanchez-Valle has received consulting fees from Wave Pharmaceuticals and Ionis-Biogen and from Roche Diagnostics and Janssen for educational activities and is member of the Data Safety Monitoring Board for Wave Pharmaceuticals and Ionis-Biogen. B. Borroni has received consulting fees from Alector and Wave Pharmaceuticals and has a pending patent on noninvasive brain stimulation. M. Masellis has received compensation for royalties from Henry Stewart Talks Ltd.; consulting fees from Arkuda Therapeutics, Ionis, Alector, Biogen, and Wave Life Sciences; personal fees for educational activities from Alector and Arkuda Therapeutics; and travel fees from Alector Pharmaceuticals. M.C. Tartaglia has received consulting fees from Roche. C. Graff has received consulting fees from Studentlitteratur 238 SEK 2020 and reimbursement for travel and lodging from University of Pennsylvania for attending KOL-meeting in October 2019. J. Rowe has received consulting fees from Asceneuron, Biogen, UCB, SV Health, and Astex; has provided expert testimony in private noncommercial medicolegal case reports; is part of an Advisory Board for several noncommercial academic institutions; and is Chief Scientific Advisor to Alzheimers Research UK, Guarantor of Brain, Trustee PSP Association, Trustee Darwin College, and Associate Director of the Dementias Platform UK. E. Finger has received consulting fees for the AAN annual meeting speaker and course director honorariums; is member of the Data Safety Monitoring Board for the Lithium trial, PI E. Huey (trial funded by peer reviewed grants from ADDF); and is a scientific advisory board member for Vigil Neuroscience, Denali Therapeutics, and on an advisory panel for Biogen. M. Synofzik has received consulting fees from Janssen Pharmaceuticals, Ionis Pharmaceuticals, and Orphazyme Pharmaceuticals and received support from the Movement Disorder Society for travel. R. Vandenberghe has received consulting fees from CyTox and is a member of the Data Safety and Monitoring Board of AC Immune. I. Santana has received consulting fees from Biogen and Roche Biogen, personal fees for presentations from Biogen, and is part of the Data Safety Monitoring Board for Novo Nordisk. S. Ducharme has received consulting fees from Innodem Neurosciences and personal fees from Sunovion Eisai. J. Levin has received consulting fees from Bayer Vital, Roche, and Biogen; is part of the Advisory Board for Axon Neurosciences; has received compensation for duty as part-time CMO from Modag; and has received author fees from Thieme medical publishers and W. Kohlhammer GmbH medical publishers as well as nonfinancial support from AbbVie. M. Otto has received consulting fees from Biogen, Axon, and Roche and is part of the Advisory Board for Axon. I. Le Ber has received consulting fees from Alector Prevail Therapeutics and personal fees from MDS and is also part of the Data Safety Monitoring Board for Alector Prevail Therapeutics. K. Kentarci served on the Data Safety Monitoring Board for Takeda Global Research & Development Center and data monitoring boards of Pfizer and Janssen Alzheimer Immunotherapy and received research support from Avid Radiopharmaceuticals, Eli Lilly, the Alzheimer’s Drug Discovery Foundation and the NIH. B.F. Boeve has served as an investigator for clinical trials sponsored by Alector, Biogen, Transposon and Cognition Therapeutics; he serves on the Scientific Advisory Board of the Tau Consortium which is funded by the Rainwater Charitable Foundation. A.L. Boxer has served as a consultant for Aeovian, AGTC, Alector, Arkuda, Arvinas, Boehringer Ingelheim, Denali, GSK, Life Edit, Humana, Oligomerix, Oscotec, Roche, TrueBinding, Wave, Merck and received research support from Biogen, Eisai and Regeneron. H.J. Rosen has received research support from Biogen Pharmaceuticals, has consulting agreements with Wave Neuroscience, Ionis Pharmaceuticals, Eisai Pharmaceuticals, and Genentech.

## APPENDIX 2: Co-investigators

Co-investigators in the GENFI and ALLFTD consortia are listed in the eAppendix.

## Notes

### Author Declarations

This study was reviewed and approved by the human research ethics board at Western University, London, Canada, and by the local ethics committees at each of the Frontotemporal Dementia Prevention Initiative sites, as stated in the Methods section of our manuscript.

