## Supplementary material for "Neurodevelopmental effects of genetic frontotemporal dementia mutations revealed by total intracranial volume differences": eAppendix

#### GENFI Investigators

Martina Bocchetta,[1] David Cash,[1] Rhian Convery,[1] Sophie Goldsmith,[1] Kiran Samra,[1] David L. Thomas,[2] Thomas Cope,[3] Maura Malpetti,[4] Antonella Alberici,[5] Enrico Premi,[6] Roberto Gasparotti,[7] Emanuele Buratti,[8] Valentina Cantoni,[9] Andrea Arighi,[10] Chiara Fenoglio,[10,11] Vittoria Borracci,[11] Maria Serpente,[11] Tiziana Carandini,[11] Emanuela Rotondo,[11] Giacomina Rossi,[12] Giorgio Giaccone,[12] Giuseppe Di Fede,[12] Paola Caroppo,[12] Sara Prioni,[12] Veronica Redaelli,[12] David Tang-Wai,[13] Ekaterina Rogaeva,[14] Johanna Krüger,[15] Miguel Castelo-Branco,[16] Morris Freedman,[17] Ron Keren,[18] Sandra Black,[19] Sara Mitchell,[19] Christen Shoesmith,[20] Robert Bartha,[21] Rosa Rademakers,[22] Jackie Poos,[23] Janne M. Papma,[23] Lucia Giannini,[23] Liset de Boer,[23] Julie de Houwer,[23] Rick van Minkelen,[24] Yolande Pijnenburg,[25] Benedetta Nacmias,[26] Camilla Ferrari,[26] Cristina Polito,[27] Gemma Lombardi,[28] Valentina Bessi,[28] Enrico Fainardi,[29] Stefano Chiti,[29] Mattias Nilsson,[30] Henrik Viklund,[31] Melissa Taheri Rydell,[32] Vesna Jelic,[33] Linn Öijerstedt,[34] Tobias Langheinrich,[34] Albert Lladó,[35] Anna Antonell,[35] Jaume Olives,[35] Mircea Balasa,[35] Nuria Bargalló,[36] Sergi Borrego-Ecija,[35] Ana Verdelho,[37] Carolina Maruta,[38] Tiago Costa-Coelho,[39] Gabriel Miltenberger,[39] Frederico Simões do Couto,[40] Alazne Gabilondo,[41] Ioana Croitoru,[42] Mikel Tainta,[42] Myriam Barandiaran,[43] Patricia Alves,[44] Benjamin Bender,[45] David Mengel,[46] Lisa Graf,[46] Annick Vogels,[47] Mathieu Vandenbulcke,[48] Philip Van Damme,[49] Rose Bruffaerts,[50] Koen Poesen,[51] Pedro Rosa-Neto,[52] Maxime Montembault,[53] Raphaella Lara Migliaccio,[54] Ninon Burgos,[54] Daisy Rinaldi,[54] Catharina Prix,[55] Elisabeth Wlasich,[55] Olivia Wagemann,[55] Sonja Schönecker,[55] Alexander Maximilian Bernhardt,[55] Anna Stockbauer,[55] Jolina Lombardi,[56] Sarah Anderl-Straub,[56] Adeline Rollin,[57] Gregory Kuchcinski,[58] Maxime Bertoux,[59] Thibaud Lebouvier,[58] Vincent Deramecourt,[58] João Durães,[60] Marisa Lima,[60] Maria João Leitão,[61] Maria Rosario Almeida,[62] Miguel Tábuas-Pereira,[60,62] Sónia Afonso,[63] João Lemos[62]

1 Department of Neurodegenerative Disease, Dementia Research Centre, UCL Queen Square Institute of Neurology, London, UK
2 Neuroimaging Analysis Centre, Department of Brain Repair and Rehabilitation, UCL Institute of Neurology, Queen Square, London, UK
3 Cambridge University Hospitals NHS Trust, Cambridge UK
4 Department of Clinical Neurosciences, University of Cambridge, Cambridge, UK
5 Centre for Neurodegenerative Disorders, Department of Clinical and Experimental Sciences, University of Brescia, Brescia, Italy
6 Stroke Unit, ASST Brescia Hospital, Brescia, Italy
7 Neuroradiology Unit, University of Brescia, Brescia, Italy
8 ICGEB Trieste, Italy
9 Centre for Neurodegenerative Disorders, Department of Clinical and Experimental Sciences, University of Brescia, Brescia, Italy
10 Fondazione IRCCS Ca' Granda Ospedale Maggiore Policlinico, Neurodegenerative Diseases Unit, Milan, Italy
11 University of Milan, Centro Dino Ferrari, Milan, Italy

12 Fondazione IRCCS Istituto Neurologico Carlo Besta, Milano, Italy
13 The University Health Network, Krembil Research Institute, Toronto, Canada
14 Tanz Centre for Research in Neurodegenerative Diseases, University of Toronto, Toronto, Canada
15 Research Unit of Clinical Medicine, Neurology, University of Oulu, Oulu, Finland. Neurocenter, Neurology, Oulu University Hospital, Oulu, Finland.
16 Faculty of Medicine, ICNAS, CIBIT, University of Coimbra, Coimbra, Portugal.
17 Baycrest Health Sciences, Rotman Research Institute, University of Toronto, Toronto, Canada
18 The University Health Network, Toronto Rehabilitation Institute, Toronto, Canada
19 Sunnybrook Health Sciences Centre, Sunnybrook Research Institute, University of Toronto, Toronto, Canada
20 Department of Clinical Neurological Sciences, University of Western Ontario, London, Ontario, Canada
21 Department of Medical Biophysics, The University of Western Ontario, London, Ontario, Canada. Centre for Functional and Metabolic Mapping, Robarts Research Institute, The University of Western Ontario, London, Ontario, Canada.
22 Center for Molecular Neurology, University of Antwerp
23 Department of Neurology, Erasmus Medical Center, Rotterdam, Netherlands
24 Department of Clinical Genetics, Erasmus Medical Center, Rotterdam, Netherlands
25 Amsterdam University Medical Centre, Amsterdam VUMC, Amsterdam, Netherlands
26 Department of Neuroscience, Psychology, Drug Research and Child Health, University of Florence, Florence, Italy
27 Department of Biomedical, Experimental and Clinical Sciences "Mario Serio", Nuclear Medicine Unit, University of Florence, Florence, Italy
28 Department of Neuroscience, Psychology, Drug Research and Child Health, University of Florence, Florence, Italy
29 Neuroradiology Unit, Department of Experimental and Clinical Biomedical Sciences, University of Florence, Florence, Italy
30 Department of Clinical Neuroscience, Karolinska Institutet, Stockholm, Sweden
31 Karolinska University Hospital Huddinge
32 Department of Neurobiology, Care Sciences and Society; Center for Alzheimer Research, Division of Neurogeriatrics, Bioclinicum, Karolinska Institutet, Solna, Sweden. Unit for Hereditary Dementias, Theme inflammation and Aging, Karolinska University Hospital, Solna, Sweden.
33 Department of Neurobiology, Care Sciences and Society; Division of Clinical Geriatrics, Karolinska Institutet, Stockholm, Sweden. Cognitive clinic, Theme inflammation and Aging, Karolinska University Hospital, Solna, Sweden.
34 Division of Neuroscience and Experimental Psychology, Wolfson Molecular Imaging Centre, University of Manchester, Manchester, UK. Manchester Centre for Clinical Neurosciences, Department of Neurology, Salford Royal NHS Foundation Trust, Manchester, UK.
35 Alzheimer's disease and Other Cognitive Disorders Unit, Neurology Service, Hospital Clínic, Barcelona, Spain
36 Imaging Diagnostic Center, Hospital Clínic, Barcelona, Spain

37 Department of Neurosciences and Mental Health, Centro Hospitalar Lisboa Norte - Hospital de Santa Maria & Faculty of Medicine, University of Lisbon, Lisbon, Portugal
38 Laboratory of Language Research, Centro de Estudos Egas Moniz, Faculty of Medicine, University of Lisbon, Lisbon, Portugal
39 Faculty of Medicine, University of Lisbon, Lisbon, Portugal
40 Faculdade de Medicina, Universidade Católica Portuguesa
41 Cognitive Disorders Unit, Department of Neurology, Donostia University Hospital, San Sebastian, Gipuzkoa, Spain
42 Instituto de Investigación Sanitaria Biogipuzkoa, Neurosciences Area, Group of Neurodegenerative Diseases, San Sebastian, Spain.
43 Cognitive Disorders Unit, Department of Neurology, Donostia University Hospital, San Sebastian, Gipuzkoa, Spain
44 Instituto de Investigación Sanitaria Biogipuzkoa, Neurosciences Area, Group of Neurodegenerative Diseases, San Sebastian, Spain.
45 Department of Diagnostic and Interventional Neuroradiology, University of Tübingen, Tübingen, Germany
46 Department of Neurodegenerative Diseases, Hertie-Institute for Clinical Brain Research and Center of Neurology, University of Tübingen, Tübingen, Germany
47 Department of Human Genetics, KU Leuven, Leuven, Belgium
48 Geriatric Psychiatry Service, University Hospitals Leuven, Belgium; Neuropsychiatry, Department of Neurosciences, KU Leuven, Leuven, Belgium
49 Neurology Service, University Hospitals Leuven, Belgium; Laboratory for Neurobiology, VIB-KU Leuven Centre for Brain Research, Leuven, Belgium
50 Department of Biomedical Sciences, University of Antwerp, Antwerp, Belgium; Biomedical Research Institute, Hasselt University, 3500 Hasselt, Belgium
51 Laboratory for Molecular Neurobiomarker Research, KU Leuven, Leuven, Belgium
52 Translational Neuroimaging Laboratory, McGill Centre for Studies in Aging, McGill University, Montreal, Québec, Canada
53 Douglas Research Centre, Department of Psychiatry, McGill University, Montreal, Québec, Canada
54 Sorbonne Université, Paris Brain Institute – Institut du Cerveau – ICM, Inserm U1127, CNRS UMR 7225, AP-HP - Hôpital Pitié-Salpêtrière, Paris, France
55 Neurologische Klinik, Ludwig-Maximilians-Universität München, Munich, Germany
56 Department of Neurology, University of Ulm, Ulm, Germany
57 CHU, CNR-MAJ, Labex Distalz, LiCEND Lille, France
58 Univ Lille, France; Inserm 1172, Lille, France; CHU, CNR-MAJ, Labex Distalz, LiCEND Lille, France
59 Inserm 1172, Lille, France; CHU, CNR-MAJ, Labex Distalz, LiCEND Lille, France
60 Neurology Department, Centro Hospitalar e Universitario de Coimbra, Coimbra, Portugal
61 Centre of Neurosciences and Cell Biology, Universidade de Coimbra, Coimbra, Portugal
62 Faculty of Medicine, University of Coimbra, Coimbra, Portugal
63 Instituto Ciencias Nucleares Aplicadas a Saude, Universidade de Coimbra, Coimbra, Portugal

#### ALLFTD Investigators

Liana Apostolova,[1] Brian Appleby,[2] Sami Barmada,[3] Bradley Boeve,[4] Yvette Bordelon,[5] Hugo Botha,[4] Adam L. Boxer,[6] Andrea Bozoki,[7] Danielle Brushaber,[8] David Clark,[1] Giovanni Coppola,[5] Ryan Darby,[9] Bradford C. Dickerson,[10] Dennis Dickson,[11] Kimiko Domoto-Reilly,[12] Kelley Faber,[13] Anne Fagan,[14] Julie A. Fields,[15] Tatiana Foroud,[16] Leah Forsberg,[4] Daniel Geschwind,[5,17] Nupur Ghoshal,[14] Jill Goldman,[18] Douglas R. Galasko,[19] Ralitza Gavrilova,[4] Tania F. Gendron,[11] Jonathon Graff-Radford,[4] Neill Graff-Radford,[20] Ian M. Grant,[21] Chadwick Hales,[22] Matthew G. H. Hall,[6] Eric Huang,[6] Hilary W. Heuer,[6] Lawrence Honig,[23] Ging-Yuek Hsiung,[24] Edward D. Huey,[23] David Irwin,[24] Kejal Kantarci,[4] Daniel Kaufer,[26] Diana Kerwin,[27] David Knopman,[4] John Kornak,[28] Joel Kramer,[6] Walter Kremers,[8] Justin Kwan,[29] Maria Lapid,[4] Argentina Lario Lago,[6] Suzee Lee,[6] Gabriel Leger,[19] Peter Ljubenkov,[6] Irene Litvan,[30] Diane Lucente,[10] Ian R. Mackenzie,[30] Joseph C. Masdeux,[31] Scott McGinnis,[10] Mario Mendez,[5] Carly Mester,[8] Bruce L. Miller,[6] Chiadi Onyike,[32] M. Belen Pascual,[31] Leonard Petrucelli,[11] Peter Pressman,[33] Rosa Rademakers,[11] Vijay Ramanan,[4] Eliana Marisa Ramos,[5] Katya Rascovsky,[25] Katherine P. Rankin,[6] Aaron Ritter,[33] Erik Roberson,[34] Julio C. Rojas,[6] Howard J. Rosen,[6] Rodolfo Savica,[4] William W. Seeley,[6] Allison Snyder,[29] Adam M. Staffaroni,[6] Anna Campbell Sullivan,[35] Jeremy Syrjanen,[4] M. Carmela Tartaglia,[36] Jack C. Taylor,[6] Lawren VandeVrede,[6] Sandra Weintraub,[37] Dylan Wint,[38] and Bonnie Wong[10]

1 Department of Neurology, Indiana University, Indianapolis, IN, USA
2 Department of Neurology, Case Western Reserve University, Cleveland, OH, USA
3 Department of Neurology, University of Michigan, Ann Arbor, MI, USA
4 Department of Neurology, Mayo Clinic, Rochester, MN, USA
5 Department of Neurology, University of California, Los Angeles, Los Angeles, CA, USA
6 Department of Neurology, Memory and Aging Center, Weill Institute for Neurosciences, University of California, San Francisco, San Francisco, CA, USA
7 Department of Neurology, University of North Carolina, Chapel Hill, NC, USA
8 Department of Quantitative Health Sciences, Mayo Clinic, Rochester, MN, USA
9 Department of Neurology, Vanderbilt University, Nashville, TN, USA
10 Department of Neurology, Massachusetts General Hospital and Harvard Medical School, Boston, MA, USA
11 Department of Neuroscience, Mayo Clinic, Jacksonville, FL, USA
12 Department of Neurology, University of Washington, Seattle, WA, USA
13 Indiana University School of Medicine, National Centralized Repository for Alzheimer's, Indianapolis, IN, USA
14 Departments of Neurology and Psychiatry, Washington University School of Medicine, Washington University, St. Louis, MO, USA
15 Department of Psychiatry and Psychology, Mayo Clinic, Rochester, MN, USA
16 Indiana University School of Medicine, National Centralized Repository for Alzheimer's, Indianapolis, IN, USA
17 Institute for Precision Health, David Geffen School of Medicine, University of California, Los Angeles, Los Angeles, CA, USA
18 Department of Neurology, Columbia University, New York, NY, USA
19 Department of Neurosciences, University of California, San Diego, La Jolla, CA, USA

20 Department of Neurology, Mayo Clinic, Jacksonville, FL, USA.
21 Department of Neurology, Northwestern University, Chicago, IL, USA.
22 Department of Neurology, Emory University, Atlanta, GA, USA
23 Department of Neurology, Columbia University, New York, NY, USA
24 Division of Neurology, University of British Columbia, Vancouver, British Columbia, Canada
25 Department of Neurology, University of Pennsylvania, Philadelphia, PA, USA
26 Department of Neurology, University of North Carolina, Chapel Hill, NC, USA
27 Department of Neurology, UT Southwestern, Dallas, TX, USA
28 Department of Epidemiology and Biostatistics, University of California, San Francisco, San Francisco, CA, USA
29 Neurodegeneration Disorders Clinic, National Institute of Neurological Disorders and Stroke, Bethesda, MD, USA
30 Department of Pathology, University of British Columbia, Vancouver, British Columbia, Canada
31 Department of Neurology, Houston Methodist, Houston, TX, USA
32 Department of Psychiatry and Behavioral Sciences, Johns Hopkins University, Baltimore, MD, USA
33 Department of Neurology, University of Colorado, Aurora, CO, USA
34 Department of Neurology, University of Alabama at Birmingham, Birmingham, AL, USA
35 Department of Neurology, University of Texas Health Science Center at San Antonio, San Antonio, TX, USA
36 Tanz Centre for Research in Neurodegenerative Diseases, Division of Neurology, University of Toronto, Toronto, Ontario, Canada
37 Department of Biomedical Sciences, University of Antwerp, Antwerp, Belgium
38 Lou Ruvo Center for Brain Health, Cleveland Clinic, Nevada, Las Vegas, NV, USA

### Participant Demographics by Cohort

#### GENFI cohort

A total of 924 participants were included in the GENFI cohort analysis, which comprised 595 mutation carriers and 329 non-carriers (eTable 1a). Within the GENFI cohort, there were 382 symptomatic carriers, 213 pre-symptomatic carriers, 329 non-carriers. Participants were further divided by genetic mutation, resulting in 228 *GRN* carriers, 145 *GRN* familial non-carriers, 96 *MAPT* carriers, 58 *MAPT* familial non-carriers, 271 *C9orf72* repeat expansion carriers, and 126 *C9orf72* familial non-carriers. The mean age of all participants at their baseline visit was 50.7 years (SD=12.4, range=23.9 to 85.7).

There were also no significant differences in age, birth decade, sex, race, and handedness between carriers and non-carriers for *GRN* and *MAPT* ($p$>0.05).*C9orf72* carriers differed significantly with their non-carrier counterparts in age ($p$=2.04e-5, $d$=0.46) and birth decade ($p$=3.02e5, $d$=-0.41), with carriers being older and born in earlier decades; however, these differences were all small in effect size ($d$<0.5). No significant differences were observed between *C9orf72* carriers and non-carriers in sex, race, and handedness.

#### ALLFTD cohort

A total of 510 participants were examined in the ALLFTD cohort, of whom 307 were mutation carriers, and 203 were familial non-carriers (eTable 1b). This was further categorized into 70 *GRN* carriers, 56 *GRN* familial non-carriers, 91 *MAPT* carriers, 56 *MAPT* familial non-carriers, 146 *C9orf72* repeat expansion carriers, and 91 *C9orf72* familial non-carriers. Participants had a mean age of 47.9 years (SD of 14.3, range of 18 to 79). Carriers and non-carriers per genetic mutation did not differ significantly in the demographic characteristics of age, birth decade, sex, nor handedness ($p$>0.05).

### Covariates Significantly Associated with Outcome Variable in *GRN* Models Computed for Sensitivity Analysis

#### *GRN* TIV, birth decade sensitivity analysis

Sensitivity analysis with birth decade as an interaction term revealed that it did not predict the main effect of TIV differences between *GRN* carriers and non-carriers, while controlling for sex and site (p>0.05).

#### *GRN* TIV, sex sensitivity analysis

There was a trend of larger TIV in male *GRN* mutation carriers compared to non-carriers: $F$(1,210)=3.20, $p$=0.08, $\eta_p^2$=0.02. Birth decade ($F$(7,210)=3.18, $p$=0.003, $\eta_p^2$=0.11) and site ($F$(26,210)=2.78, $p$=3.92e-5, $\eta_p^2$=0.29) were significant main effects.

No significant difference was observed in the female *GRN* model, although the means are trending in the same direction as the primary analysis and that of the male *GRN* model, with mean TIV of female *GRN* carriers larger than non-carriers: $F(1,282)=0.23$, $p=0.63$, $\eta_p^2=0.0009$. Site was a significant main effect: $F(27,282)=2.11$, $p=0.002$, $\eta_p^2=0.19$.

##### *GRN* TIV, site sensitivity analysis

Findings from the site sensitivity analysis, where sites containing only one participant was removed, were consistent with that of the primary analysis: $F(1,491)=3.89$, $p=0.049$, $\eta_p^2=0.008$. When the GENFI and ALLFTD cohort sites were analyzed separately, significance was observed only in the ALLFTD cohort ($F(1,123)=4.11$, $p=0.045$, $\eta_p^2=0.04$), but not in the GENFI cohort ($F(1,370)=1.04$, $p=0.31$, $\eta_p^2=0.003$).

In the GENFI cohort analysis, sex ($F(1,370)=228.8$, $p<2.2e-16$, $\eta_p^2=0.40$) and site ($F(17,370)=2.86$, $p=1.50e-4$, $\eta_p^2=0.12$) differences were significant associated with TIV. In particular, males were associated with having TIV. In the ALLFTD cohort analysis, sex was associated with TIV changes, with males having larger TIV than females: $F(1,123)=121.4$, $p<2e-16$, $\eta_p^2=0.54$.

##### *GRN* education, site sensitivity analysis

When sites containing only one participant was removed, the results were consistent with that of the primary analysis ($p>0.05$). In the GENFI cohort model, birth decade and site were significantly associated with education: $F(7,371)=5.96$, $p=1.45e-6$, $\eta_p^2=0.11$, and $F(17,371)=5.69$, $p=2.01e-11$, $\eta_p^2=0.22$, respectively. There were no significant main effects in the ALLFTD cohort model.

#### Covariates Significantly Associated with Outcome Variable in *MAPT* Models Computed for Sensitivity Analysis

##### *MAPT* TIV, sex sensitivity analysis

In the male *MAPT* model for TIV, site ($F(22,132)=1.98$, $p=0.01$, $\eta_p^2=0.29$) was significantly associated with TIV. There were no significant main effects in the female *MAPT* model for TIV.

##### *MAPT* education, site sensitivity analysis

When sites containing only one participant was removed, the results were consistent with that of the primary analysis ($p>0.05$). In the GENFI cohort analysis, where *MAPT* carriers trended towards having fewer years of education than non-carriers (as reported in the main manuscript), birth decade ($F(5,152)=2.13$, $p=0.07$, $\eta_p^2=0.08$) and site ($F(15,152)=1.56$, $p=0.09$, $\eta_p^2=0.15$) trended towards significance in relation to education. In the ALLFTD cohort analysis, there were no significant main effects.

#### Covariates Significantly Associated with Outcome Variable in *C9orf72* Models Computed for Sensitivity Analysis

##### *C9orf72* TIV, sex sensitivity analysis

In the female and male *C9orf72* models, site was correlated to TIV: $F(38,345)=2.27$, $p=7.62e-5$, $\eta_p^2=0.22$, and $F(35,278)=1.91$, $p=0.003$, $\eta_p^2=0.22$, respectively. In the female *C9orf72* model, birth decade showed a trend towards significance in its correlation to TIV: $F(6,345)=1.84$, $p=0.09$, $\eta_p^2=0.04$ with those being born in later decades having larger TIV.

##### *C9orf72* TIV, site sensitivity analysis

When sites containing only one participant was removed, the results were consistent with that of the primary analysis ($p>0.05$). When GENFI and ALLFTD cohorts were analyzed separately, covariates were related to TIV in the GENFI cohort analysis; sex: $F(1,394)=348.3$, $p<2.2e-16$, $\eta_p^2=0.49$; site: $F(24,394)=1.91$, $p=0.007$, $\eta_p^2=0.11$; and birth decade showed a trend towards significance in its association to TIV: $F(6,394)=1.84$, $p=0.09$, $\eta_p^2=0.03$. In the ALLFTD cohort analysis of *C9orf72* participants, sex was associated with TIV, where males had greater TIV than females: $F(1,233)=140.9$, $p<2e-16$, $\eta_p^2=0.40$.

##### *C9orf72* education, site sensitivity analysis

When sites containing only one participant was removed, the results were consistent with that of the primary analysis ($p>0.05$). In the GENFI cohort analysis of *C9orf72* participants, birth decade ($F(6,395)=3.84$, $p=0.001$, $\eta_p^2=0.06$) and site ($F(24,395)=3.80$, $p=1.46e-8$, $\eta_p^2=0.20$) were significantly related to years of education. Sex was found to trend towards significance in its association with education: $F(1,395)=3.04$, $p=0.08$, $\eta_p^2=0.008$. In the ALLFTD cohort analysis, site was related to education: $F(16,234)=3.76$, $p=3.99e-6$, $\eta_p^2=0.22$.

#### eTable 1a. Demographic characteristics of participants in the GENFI cohort

| Characteristic | All | | | *GRN* | | | *MAPT* | | | *C9orf72* | | |
|---|---|---|---|---|---|---|---|---|---|---|---|---|
| | Total | Carriers | Non-carriers | Carriers | Non-carriers | Between group comparison | Carriers | Non-carriers | Between group comparison | Carriers | Non-carriers | Between group comparison |
| Sample size (*n*) | 924 | 595 | 329 | 228 | 145 | NA | 96 | 58 | NA | 271 | 126 | NA |
| Age, years (mean ± SD) | 50.7 ± 12.4 | 52.0 ± 12.5 | 48.3 ± 11.9 | 52.6 ± 11.9 | 50.1 ± 12.9 | *t*=1.79, *p*=0.07, *d*=0.19 | 47.6 ± 12.0 | 46.3 ± 10.0 | *t*=0.64, *p*=0.52, *d*=0.34 | 53.1 ± 12.9 | 47.3 ± 11.2 | *t*=4.31, *p*= 2.04e-5***, *d*=0.46 |
| Birth Decade (*n*) | | | | | | | | | | | | |
| 1920-1929 | 2 | 0 | 2 | 0 | 2 | | 0 | 0 | | 0 | 0 | |
| 1930-1939 | 16 | 10 | 6 | 4 | 4 | | 0 | 0 | | 6 | 2 | |
| 1940-1949 | 105 | 78 | 27 | 31 | 13 | $x^2$=12.8, | 7 | 4 | $x^2$=7.86, | 40 | 10 | $x^2$=30.6, *p*= |
| 1950-1959 | 202 | 144 | 58 | 57 | 34 | *p*=0.08, | 20 | 7 | *p*=0.16, | 67 | 17 | 3.02e-5***, |
| 1960-1969 | 205 | 145 | 60 | 61 | 28 | *d*=-0.12 | 21 | 11 | *d*=-0.10 | 63 | 21 | *d*=-0.41 |
| 1970-1979 | 269 | 135 | 134 | 53 | 49 | | 28 | 29 | | 54 | 56 | |
| 1980-1989 | 118 | 80 | 38 | 21 | 13 | | 19 | 7 | | 40 | 18 | |
| 1990-1999 | 7 | 3 | 4 | 1 | 2 | | 1 | 0 | | 1 | 2 | |
| Sex (*n*) female: male | 509:415 | 321:274 | 188:141 | 136:92 | 84:61 | $x^2$=0.05, *p*=0.83, *d*=-0.03 | 51:45 | 31:27 | $x^2$=0, *p*=1, *d*=0.006 | 134:137 | 73:53 | $x^2$=2.16, *p*=0.14, *d*=0.17 |
| Education, years (mean ± SD) | 14.0 ± 3.63 | 13.7 ± 3.62 | 14.4 ± 3.61 | 13.8 ± 3.97 | 14.3 ± 4.01 | NA | 13.8 ± 3.43 | 14.8 ± 3.39 | NA | 13.6 ± 3.38 | 14.3 ± 3.21 | NA |
| Race (*n*) | | | | | | | | | $x^2$=4.72, | | | $x^2$=3.08, |
| White | 915 | 588 | 327 | 228 | 145 | NA | 91 | 57 | *p*=0.19, | 269 | 125 | *p*=0.38, |
| Non-White | 9 | 7 | 2 | 0 | 0 | | 5 | 1 | *d*=0.12 | 2 | 1 | *d*=0.06 |
| Handedness (*n*) | | | | | | | | | | | | |
| Right | 849 | 544 | 305 | 207 | 139 | $x^2$=4.60, | 88 | 52 | $x^2$=0.02, | 249 | 114 | $x^2$=0.97, |
| Left | 64 | 44 | 20 | 18 | 5 | *p*=0.10, | 8 | 6 | *p*=0.90, | 18 | 9 | *p*=0.62, |
| Ambidextrous | 10 | 6 | 4 | 3 | 1 | *d*=0.21 | 0 | 0 | *d*=-0.07 | 3 | 3 | *d*=-0.09 |
| Unknown | 0 | 0 | 0 | 0 | 0 | | 0 | 0 | | 0 | 0 | |
| Blinded Sites (*n*) | 25 | 25 | 22 | 17 | 15 | NA | 16 | 10 | NA | 25 | 21 | NA |

*p<0.05, ***p<0.001 between carriers and non-carriers. Blinded sites (*n*) refer to the number of sites which saw the specified group of participants. Race was collapsed into “white” and “non-white” to protect identities of the few who identified as non-white. An independent t-test was not performed between groups for variable of education, since this was a planned secondary outcome that would be analyzed in greater detail. Abbreviations: SD, standard deviation; NA, not applicable.

#### eTable 1b. Demographic characteristics of participants in the ALLFTD cohort

| Characteristic | All | | | *GRN* | | | *MAPT* | | | *C9orf72* | | |
|---|---|---|---|---|---|---|---|---|---|---|---|---|
| | Total | Carriers | Non-carriers | Carriers | Non-carriers | Between group comparison | Carriers | Non-carriers | Between group comparison | Carriers | Non-carriers | Between group comparison |
| Sample size (*n*) | 510 | 307 | 203 | 70 | 56 | NA | 91 | 56 | NA | 146 | 91 | NA |
| Age, years (mean ± SD) | 47.9 ± 14.3 | 48.8 ± 14.3 | 46.5 ± 14.2 | 55.5 ± 14.1 | 51.0 ± 13.6 | *t*=1.82, *p*=0.07, *d*=0.33 | 43.7 ± 13.1 | 40.9 ± 14.4 | *t*=1.22, *p*=0.23, *d*=0.21 | 48.9 ± 13.9 | 47.2 ± 13.4 | *t*=0.92, *p*=0.36, *d*=0.12 |
| Birth Decade (*n*) | | | | | | $x^2$=4.90, *p*=0.43, *d*=-0.29 | | | $x^2$=4.06, *p*=0.67, *d*=-0.18 | | | $x^2$=1.74, *p*=0.88, *d*=-0.02 |
| 1920-1929 | 0 | 0 | 0 | 0 | 0 | | 0 | 0 | | 0 | 0 | |
| 1930-1939 | 0 | 0 | 0 | 0 | 0 | | 0 | 0 | | 0 | 0 | |
| 1940-1949 | 42 | 27 | 15 | 12 | 4 | | 3 | 3 | | 12 | 8 | |
| 1950-1959 | 103 | 64 | 39 | 25 | 16 | | 11 | 5 | | 28 | 18 | |
| 1960-1969 | 117 | 73 | 44 | 15 | 18 | | 23 | 10 | | 35 | 16 | |
| 1970-1979 | 96 | 58 | 38 | 6 | 5 | | 23 | 11 | | 29 | 22 | |
| 1980-1989 | 108 | 59 | 49 | 8 | 9 | | 20 | 19 | | 31 | 21 | |
| 1990-1999 | 42 | 25 | 17 | 4 | 4 | | 10 | 7 | | 11 | 6 | |
| 2000-2009 | 2 | 1 | 1 | 0 | 0 | | 1 | 1 | | 0 | 0 | |
| Sex (*n*) female: male | 294:216 | 174:133 | 120:83 | 36:34 | 29:27 | $x^2$=7.22, *p*=1, *d*=0.007 | 54:37 | 30:26 | $x^2$=0.27, *p*=0.61, *d*=-0.12 | 84:62 | 61:30 | $x^2$=1.75, *p*=0.19, *d*=0.19 |
| Education, years (mean ± SD) | 15.8 ± 4.45 | 16.0 ± 5.37 | 15.6 ± 2.47 | 15.7 ± 2.75 | 15.7 ± 2.47 | NA | 16.4 ± 9.10 | 15.2 ± 2.28 | NA | 15.8 ± 2.39 | 15.8 ± 2.58 | NA |
| Handedness (*n*) | | | | | | $x^2$=1.50, *p*=0.47, *d*=-0.13 | | | $x^2$=1.57, *p*=0.21, *d*=0.26 | | | $x^2$=4.75, *p*=0.19, *d*=-0.07 |
| Right | 450 | 267 | 183 | 62 | 53 | | 85 | 48 | | 120 | 82 | |
| Left | 47 | 32 | 15 | 6 | 2 | | 6 | 8 | | 20 | 5 | |
| Ambidextrous | 12 | 7 | 5 | 2 | 1 | | 0 | 0 | | 5 | 4 | |
| Unknown | 1 | 1 | 0 | 0 | 0 | | 0 | 0 | | 1 | 0 | |
| Blinded Sites (*n*) | 20 | 18 | 14 | 9 | 11 | NA | 11 | 8 | NA | 16 | 12 | NA |

*p<0.05, ***p<0.001 between carriers and non-carriers. Blinded sites (*n*) refer to the number of sites which saw the specified group of participants. An independent t-test was not performed between groups for variable of education, since this was a planned secondary outcome that would be analyzed in greater detail. Abbreviations: SD, standard deviation; NA, not applicable.

#### eTable 2a: *MAPT* mutation grouping

| Group | Underlying Pathology and/or Functional Consequence | Mutations |
|---|---|---|
| 1 | Mutation in exons 1, 2, 9 | R5H, R5L, G55R, K257T, I260V, L226V, G272V, IVS9-5T>C, IVS9-11G>C, IVS9-10G>C, IVS9-10G>T |
| 2 | Mutations in exon/intron 10 affecting splicing | N279K, deltaK280, L284L, L284R, S285R, C291R, N296N, K298E, IVS10+3G>A, IVS10+4A>C, IVS10+11T>C, IVS10+12C>T, IVS10+12C>A, IVS10+13A>G, IVS10+14C>T, IVS10+15A>C, IVS10+16C>T, G303V, G304S, S305N, S305I, S305S |
| 3 | Mutations in exon 10 not affecting splicing | P301T, P301S, P301L |
| 4 | Mutations in exons 11-13 with non-paired helical filament (PHF)-tau pathology | L315R, L315L, K317M, S320F, P332S, G335S, G335V, Q336H, E342V, S352L, S356T, V363A, P364S, G336R, K369I, E372G, G389R (2170G>A), G389R (2170G>C), T427M |
| 5 | Mutations in exons 11-13 with PHF-tau pathology | V337M, R406W |

This grouping was based on the approach used by Moore et al (full reference below). Some mutations remained unsorted due to their unknown nature; participants with these mutations were excluded from the *MAPT* exploratory analysis that required mutation categorization. *Reference: Moore KM, Nicholas J, Grossman M, et al. Age at symptom onset and death and disease duration in genetic frontotemporal dementia: an international retrospective cohort study. Lancet Neurol. 2020;19:145-156. doi:10.1016/S1474-4422(19)30394-1*

#### eTable 2b: Number of participants per *MAPT* mutation group per analysis

| *MAPT* Mutation Group | Primary Analysis (GENFI & ALLFTD) | | GENFI Cohort | | ALLFTD Cohort | |
|---|---|---|---|---|---|---|
| | Carriers | Non-Carriers | Carriers | Non-Carriers | Carriers | Non-Carriers |
| 1 | 10 | 2 | 5 | 1 | 5 | 1 |
| 2 | 40 | 8 | 6 | 2 | 34 | 6 |
| 3 | 68 | 47 | 37 | 31 | 31 | 16 |
| 4 | 6 | 3 | 5 | 3 | 1 | 0 |
| 5 | 34 | 13 | 14 | 5 | 20 | 8 |
| Unknown | 29 | 17 | 29 | 16 | 0 | 1 |

#### eTable 3. Summary of outcome variable means before and after covariate adjustments in the primary analysis ANCOVA models

| Model | Outcome | Carriers, non-adjusted mean ± SE [95% CI: lower CI, upper CI] | Non-Carriers, non-adjusted mean ± SE [95% CI: lower CI, upper CI] | Carriers, adjusted mean ± SE [95% CI: lower CI, upper CI] | Non-Carriers, adjusted mean ± SE [95% CI: lower CI, upper CI] | *p* | $\eta_p^2$ |
|---|---|---|---|---|---|---|---|
| *GRN* | TIV ($mm^3$) | 1439016 ± 8725.1 [1421873, 1456159] | 1432257 ± 10632.5 [1411367, 1453148] | 1444558 ± 6393.7 [1431994, 1457123] | 1424027 ± 7874.4 [1408553, 1439502] | 0.049* | 0.008 |
| *MAPT* | TIV ($mm^3$) | 1433495 ± 10502.5 [1412827, 1454163] | 1465315 ± 13451.2 [1438844, 1491787] | 1434224 ± 8331.5 [1417819, 1450628] | 1464120 ± 10940.7 [1442578, 1485662] | 0.039* | 0.016 |
| *MAPT*, with additional covariate of MAPT mutation group | | | | 1430235 ± 8715.0 [1413059, 1447412] | 1460294 ± 12271.0 [1436109, 1484479] | 0.058† | 0.016 |
| *C9orf72* | TIV ($mm^3$) | 1446301 ± 7073.8 [1432410, 1460192] | 1436230 ± 9827.9 [1416931, 1455530] | 1442488 ± 5172.2 [1432329, 1452646] | 1442823 ± 7332.4 [1428422, 1457225] | 0.97 | 2.30e6- |
| *GRN* | Education (years) | 14.3 ± 0.2 [13.8, 14.7] | 14.7 ± 0.3 [14.2, 15.2] | 14.5 ± 0.2 [14.1, 14.8] | 14.4 ± 0.2 [14.0, 14.9] | 0.85 | 8.11e-5 |
| *MAPT* | Education (years) | 14.6 ± 0.2 [14.2, 15.0] | 14.9 ± 0.3 [14.3, 15.4] | 14.6 ± 0.2 [14.1, 15.0] | 15.0 ± 0.3 [14.4, 15.5] | 0.27 | 0.005 |
| *MAPT*, with additional covariate of MAPT mutation group | | | | 14.5 ± 0.2 [14.1, 15.0] | 15.1 ± 0.3 [14.5, 15.7] | 0.15 | 0.009 |
| *C9orf72* | Education (years) | 14.5 ± 0.1 [14.3, 14.8] | 15.0 ± 0.2 [14.6, 15.4] | 14.6 ± 0.1 [14.3, 14.8] | 14.9 ± 0.2 [14.6, 15.3] | 0.14 | 0.004 |

*$p<0.05$. †$0.05<p<0.10$. Abbreviations: ANCOVA, analysis of covariance; TIV, total intracranial volume; CI, confidence interval.

#### eTable 4. Summary of outcome variable means before and after covariate adjustments in ANCOVA models computed for sensitivity analyses that were significant or trending towards significance

<table>
<tr><th>Sensitivity analysis for which covariate</th><th>Model</th><th>Outcome</th><th>Carriers, non-adjusted mean ± SE [lower CI, upper CI]</th><th>Non-Carriers, non-adjusted mean ± SE [lower CI, upper CI]</th><th>Carriers, adjusted mean ± SE [lower CI, upper CI]</th><th>Non-Carriers, adjusted mean ± SE [lower CI, upper CI]</th><th>p</th><th>$\eta_p^2$</th></tr>
<tr><td>Sex</td><td>Primary analysis: male GRN</td><td>TIV ($mm^3$)</td><td>1551651 ± 10633.7 [1530689, 1572613]</td><td>1542263 ± 12746.2 [1517136, 1567390]</td><td>1560239 ± 10053.2 [1540399, 1580078]</td><td>1529924 ± 12352.0 [1505548, 1554300]</td><td>0.075†</td><td>0.017</td></tr>
<tr><td>Sex</td><td>Primary analysis: female MAPT</td><td>TIV ($mm^3$)</td><td>1346158 ± 10317.3 [1325785, 1366531]</td><td>1385894 ± 13471.5 [1359293, 1412495]</td><td>1345118 ± 10947.1 [1323464, 1366773]</td><td>1387667 ± 14717.6 [1358554, 1416780]</td><td>0.029*</td><td>0.036</td></tr>
<tr><td>Site</td><td>GENFI cohort: MAPT</td><td>Education (years)</td><td rowspan="2">13.8 ± 0.3 [13.1, 14.4]</td><td rowspan="2">14.6 ± 0.4 [13.7, 15.5]</td><td>13.6 ± 0.3 [13.0, 14.3]</td><td>14.8 ± 0.5 [13.9, 15.7]</td><td>0.065†</td><td>0.026</td></tr>
<tr><td>Site</td><td>GENFI cohort: MAPT model, with MAPT mutation group as covariate</td><td>Education (years)</td><td>13.6 ± 0.3 [12.9, 14.3]</td><td>14.8 ± 0.5 [13.9, 15.7]</td><td>0.053†</td><td>0.029</td></tr>
<tr><td>Site</td><td>ALLFTD cohort: GRN</td><td>TIV ($mm^3$)</td><td>1504266 ± 18354.3 [1467935, 1540598]</td><td>1476856 ± 20373.7 [1436528, 1517185]</td><td>1510003 ± 12703.1 [1484815, 1535190]</td><td>1469789 ± 14221.3 [1441590, 1497987]</td><td>0.045*</td><td>0.038</td></tr>
<tr><td>Site</td><td>ALLFTD cohort: MAPT</td><td>TIV ($mm^3$)</td><td>1445700 ± 16008.0 [1414061, 1477339]</td><td>1497624 ± 20406.3 [1457292, 1537957]</td><td>1449791 ± 13073.6 [1423920, 1475661]</td><td>1490977 ± 17063.8 [1457211, 1524743]</td><td>0.069†</td><td>0.026</td></tr>
</table>

*$p<0.05$. †$0.05<p<0.10$. Only significant, or trending towards significant, findings of the main effect of group on the outcome variable are reported here. Abbreviations: ANCOVA, analysis of covariance; GENFI, genetic frontotemporal temporal dementia initiative; TIV, total intracranial volume; CI, confidence interval.

### eReferences

e1. Genetic Frontotemporal Dementia Initiative (GENFI): https://www.genfi.org/

e2. ALLFTD consortium: https://www.allftd.org/, NCT04363684

e3. Advancing Research and Treatment for Frontotemporal Lobar Degeneration study (ARTFL): NCT02365922

e4. Longitudinal Frontotemporal Lobar Degeneration study (LEFFTDS): NCT02372773
